# Intracranial aneurysm stiffness assessment using 4D flow MRI

**DOI:** 10.1101/2024.04.05.24302112

**Authors:** Satoshi Koizumi, Taichi Kin, Tetsuro Sekine, Satoshi Kiyofuji, Motoyuki Umekawa, Nobuhito Saito

**Affiliations:** Department of Neurosurgery, The University of Tokyo Hospital, Tokyo, Japan; Department of Medical Information Engineering, Graduate School of Medicine, The University of Tokyo; Department of Radiology, Nippon Medical School Musashi Kosugi Hospital

**Keywords:** 4D flow MRI, phase contrast MRI, artery stiffness, damping, intracranial aneurysm

## Abstract

**Background:** Although arterial stiffness is a known biomarker for cardiovascular events and stroke, there is limited information in the literature regarding the stiffness of intracranial aneurysms. This study aimed to assess intracranial aneurysm stiffness using four-dimensional flow magnetic resonance imaging (4D flow MRI).

**Methods:** A total of 27 aneurysms in 25 patients with internal carotid artery aneurysms were included. Using 4D flow MRI, we measured the arterial pulse waveform during a cardiac cycle at planes proximal and distal to the target aneurysm. The damping of these waveforms through the aneurysm was defined as the aneurysm damping index (ADI) and compared with that of the contralateral side. Additionally, we investigated the clinical factors associated with the ADI.

**Results:** ADI assessment was successful in all cases. The obtained ADI was 1.18 ± 0.28 (mean ± standard deviation), significantly larger than 1.0 (P=0.0027 [t-test]). The ADI on the aneurysm side was larger than on the contralateral side (1.19 ± 0.30 vs 1.05 ± 0.17, P=0.029 [t-test]). On multivariate analysis, the use of beta-blockers (β=0.46, P=0.015) and smoking history (β=-0.22, P=0.024) showed a significant correlation with ADI.

**Conclusions:** We proposed a novel method to assess the stiffness of intracranial aneurysms using 4D flow MRI. The quantitative observation of the damping of arterial pulse waves revealed a correlation between ADI and clinical factors, such as the use of antihypertensive drugs and smoking. Further studies should focus on evaluating the aneurysm stiffness and its clinical applications.

## Introduction

Arterial stiffness is a biomarker for cardiovascular events and stroke. ^1^ In studies focusing on the aorta and peripheral arteries, arterial pulse wave velocity has been used to predict the risk of cardiovascular events. ^2,3^ However, the elasticity or stiffness of cerebral arteries, frequently affected by cardiovascular diseases, has not been extensively assessed in the literature. This is mainly due to the technical challenges posed by the surrounding skull, the relatively small diameter of the cerebral arteries, and their anatomical complexities.

With the advancement of magnetic resonance imaging (MRI) technology, it is possible to non-invasively measure in vivo blood flow using magnetic resonance. Phase-contrast MRI can measure and visualize the temporal evolution of complex blood flow patterns. ^4,5^ Its application in intracranial vessels is expanding. ^6,7^ Phase-contrast MRI has demonstrated the ability to non-invasively measure the pulse wave of small intracranial arteries, including perforators. These measurements have implications in predicting cerebrovascular disease risk.^8–11^

In this study, we evaluated the feasibility of using four-dimensional (4D) flow MRI, time-resolved phase-contrast MRI that can analyze dynamic changes in blood flow during a cardiac cycle, to assess the stiffness of intracranial aneurysms. The stiffness assessment of intracranial aneurysms has scarcely been covered in the literature; hence, to quantitatively analyze stiffness, we measured the damping of the pulse wave using 4D flow MRI. Damping of flow pulsation is associated with arterial compliance, that is, the ability of elastic vessels to stretch, recoil, and absorb pulsatile energy. ^12,13^ To speculate on the role of damping in clinical application, the relationship between aneurysm damping and clinical information was analyzed.

## Methods

### Study design and participants

This study followed STROBE reporting guidelines for observational studies. This study was approved by the Institutional Review Board of our hospital (approval number 10363-[5]) and included patients with unruptured intracranial aneurysms of their internal carotid artery. Before undergoing 4D flow MRI, all participants provided informed consent. The cohort comprised 25 patients (5 male and 20 female) with 27 aneurysms, including one patient with bilateral internal carotid artery aneurysms. Another patient underwent 4D flow MRI twice, with a seven-year interval, due to a substantial increase in aneurysm size. The diameter of the aneurysms was 14.4 ± 9.0 mm, with 18 of the 27 aneurysms located in the intradural portion of the internal carotid artery. Patient and aneurysm information are summarized in **Table 1**.

**Table 1:** Patient and aneurysm characteristics summary.

|  |  |
| --- | --- |
| **Number of patients** | **25** |
| **Number of aneurysms** | **27** |
| Sex (Female) | 20 (80%) |
| Age | 64 ± 16 |
| Aneurysm diameter (mm) | 14.4 ± 9.0 |
| Extradural location | 9 (33%) |
| Hypertention | 20 (74%) |
| CCB use | 14 (52%) |
| ACEi/ARB use | 14 (52%) |
| BB use | 2 (7.4%) |
| Dyslipidemia | 13 (48%) |
| Statin use | 7 (26%) |
| Diametes mellitus | 1 (3.7%) |
| Smoking history | 12 (44%) |

Continuous variables are presented as mean ± standard deviation, while categorical data are expressed as the number of subjects (percentage of the total).
CCB: Calcium channel blocker, ACEi: Angiotensin converting enzyme inhibitor, ARB: Angiotensin receptor blocker, BB: Beta blocker.

**Table 2:** Multiple regression analysis of factors associated with PI and ADI.

| | P-value | Coefficient $\beta$ (95% CI) |
| --- | --- | --- |
| PI |  |  |
| Smoking history | 0.0032 | -0.222 (-0.355 – -0.089) |
| Systolic BP | 0.0321 | 0.0005 (0.0001 – 0.0009) |
| DWMH grading | 0.0370 | 0.043 (0.005 – 0.081) |
| ADI |  |  |
| Smoking history | 0.0239 | -0.224 (-0.406 – -0.042) |
| BB use | 0.0147 | 0.463 (0.118 – 0.878) |

ADI: Aneurysm damping index, BB: Beta blocker, BP: Blood pressure, DWMH: Deep white matter hyperintensity, PI: Pulsatility index

### MRI imaging protocols

In 22 of the 27 patients, MRI images were obtained using a 3-T scanner (SIGNA Premier; GE Healthcare, Milwaukee, WI, United States) with a 48-channel head coil. Other scanners used in the study were MAGNETOM Skyra 3T (Siemens, Erlangen, Germany) in one case and MAGNETOM Avanto 1.5T (Siemens, Erlangen, Germany) in four cases. The standard imaging parameters for 3D time-of-flight magnetic resonance angiography (TOF-MRA) were as follows: repetition time (TR), 25 ms; echo time (TE), 3.4 ms; flip angle, 20°; field of view (FOV), 20 cm; slice thickness, 0.8 mm; matrix, 512×288; voxel size, 0.4×0.7×0.8 mm; bandwidth, 122.1 Hz/pixel; and scanning time, 3 min 53 s.

For 4D flow MRI, the standard imaging parameters were as follows: TR, 5.9 ms; TE, 2.9 ms; flip angle, 15°; FOV, 20 cm; slice thickness, 1 mm; matrix size, 200 × 200; voxel size, 1.0 × 1.0 × 1.0 mm; velocity encoding, 150 cm/s; and Hyperkat, 4. Data acquisition was synchronized with the cardiac cycle using R-wave triggering. The timeframes were set to obtain 20 phases per cardiac cycle. The mean imaging duration was 12 min.

### Vessel wall extraction

The shape of the vessel wall was determined using TOF-MRA. The raw data was transferred to the image processing software, Avizo® (Thermo Fisher Scientific, Massachusetts, USA). The threshold was set using the full width at half maximum criterion. ^14^ The vessel wall was created using the surface-rendering method and used for subsequent analysis. The analysis region included vessels from the cavernous portion of the internal carotid artery (ICA) to the distal portion of the middle cerebral artery M1 and anterior cerebral artery A1.

### 4D flow MRI analysis

To minimize the spatial deviation between TOF-MRA and 4D flow MRI, we registered TOF-MRA images with the magnitude data of 4D flow MRI using affine transformation and the normalized mutual information method. The technical details of this registration were previously reported by our group. ^15^ In summary, a semi-automated algorithm was used to align the positions and postures of TOF-MRA and 4D flow MRI images. The 4D flow MRI data in the antero-posterior, right-left, and superior-inferior directions were cropped using the vessel wall and integrated into a velocity vector for each voxel at each time phase. The resulting data was transferred to the analysis software, IV-FLOW® (Maxnet Co. Ltd., Tokyo, Japan). Because some aneurysms had a fusiform shape, and it was difficult to define the neck plane of every analyzed aneurysm, we defined an inflow plane (P-in) and an outflow plane (P-out) approximately 5 mm proximal and distal to the aneurysm in the parent ICA **(Figure 1A)**.

**Figure 1:**
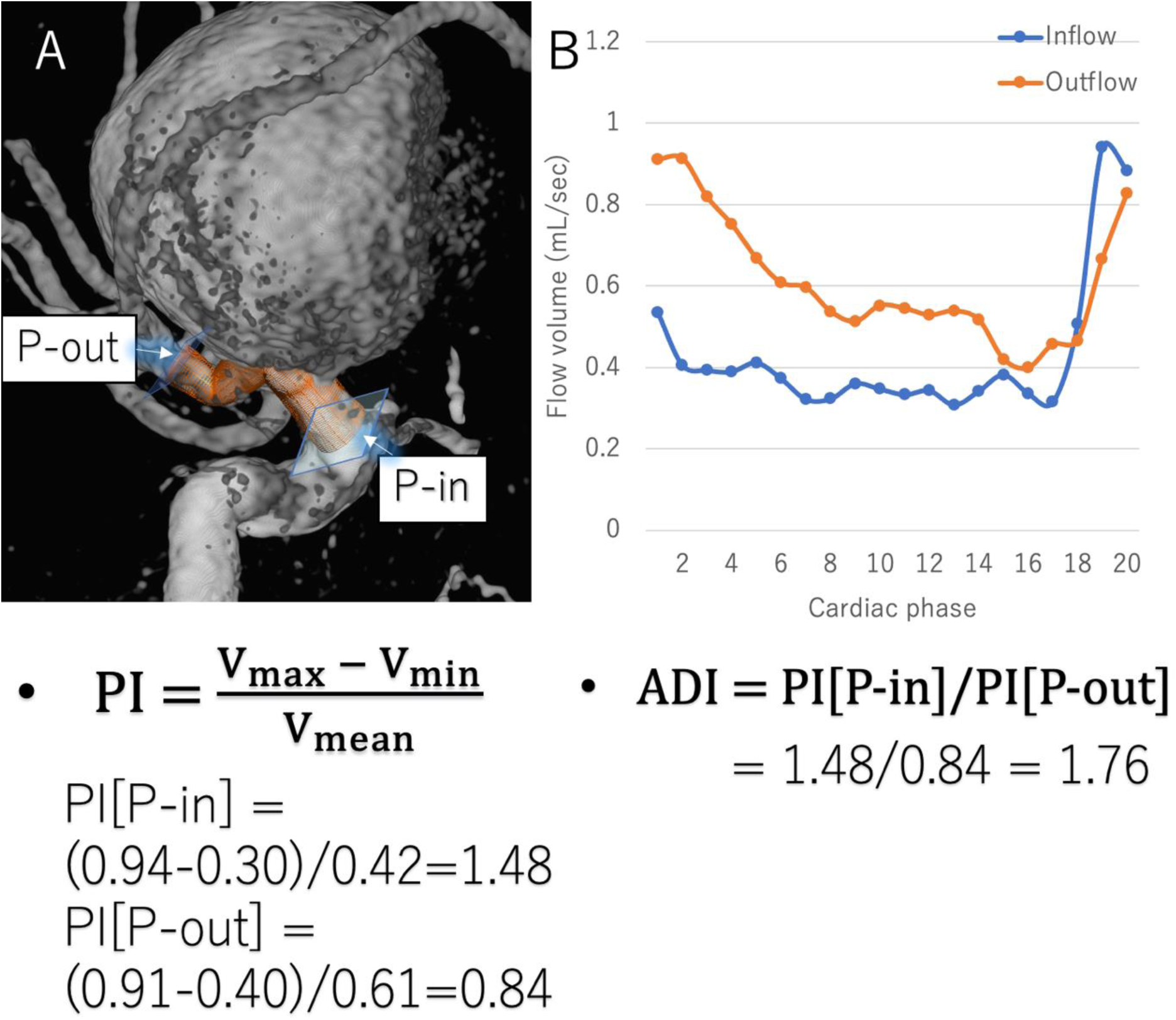
A schematic example of the aneurysm elasticity assessment method proposed in this study. 3D vessel morphology data was extracted from patients’ clinical images, and P-in and P-out were defined at the proximal and distal portions of the parent artery, respectively (Figure 1A). Flow volume waveforms were measured using 4D flow MRI at each plane, and the pulsatility index (PI) was calculated. The PI represents the extent of longitudinal waveform change during a cardiac cycle. The ratio of PI at P-in to PI at P-out is defined as the arterial damping index (ADI). If the ADI exceeds 1.0, waveform damping occurs through the aneurysm. In this case, the ADI is calculated to be 1.76 (Figure 1B).

### The elasticity assessment

For each aneurysm, flow-volume curves for P-in and P-out were generated to assess pulsatility in relation to the cardiac phase. The pulsatility index (PI) was calculated for each curve as 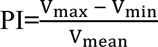, as previously described. ^8,9^ The PI increased as the flow change increased, and vice versa. However, PI directly reflects cardiac function and is unsuitable for assessing arterial wall elasticity. To quantitatively assess the elasticity of the aneurysm wall, we focused on the arterial “damping,” which refers to the attenuation of the arterial velocity curve between the proximal and distal portions of the vessel wall of interest. If the vessel wall of interest has high elasticity, the flow-volume curve of the distal region becomes flatter than that of the proximal region. If the elasticity is weak, the flow-volume curve remains uneven after passing through the region of interest. In a recent report using 7T MRI, Arts et al. quantitatively demonstrated arterial damping between middle cerebral artery M1 segment and its perforators in healthy participants. ^10^ Following their methodology, we defined aneurysm damping index (ADI) as 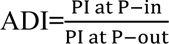 to validate if the damping through the aneurysm could be observed. If the ADI is >1, it indicates that damping of the flow volume occurs in an aneurysm. An illustrative example of PI and ADI measurements is summarized in **Figure 1B**. To compare the damping effect between a normal intracranial artery and an aneurysm, P-in and P-out of the contralateral ICA were defined at the corresponding locations of the P-in and P-out of the aneurysm side, respectively. In the 24 cases where the contralateral ICA was normal, the contralateral ADIs were also calculated and compared with those of the aneurysm side.

### Assessment of the measured elasticity in relation to the clinical information

To validate the clinical feasibility of PI and ADI, the following clinical information was collected and analyzed using a linear regression model: sex, age, aneurysm location (intra-versus extradural), aneurysm maximal diameter, deep white matter hypersensitivity grading on MRI fluid attenuation inversion recovery images, ^16^ vital signs including blood pressure and heart rate, vascular risk factors including hypertension, dyslipidemia, diabetes mellitus, and chronic kidney disease, and medications including calcium channel blockers, angiotensin-converting enzyme inhibitors, angiotensin receptor blockers, beta-blockers, diuretics and statins. White matter hyperintensity grading was performed by two board-certified neurosurgeons who were blinded to the 4D flow MRI analysis results. The inter-rater reliability of white matter hyperintensity grading was substantial between the two observers (kappa coefficient=0.782), and the sum of their grades was used in subsequent analysis.

### Statistical analysis

All statistical analyses were performed using the R software (version 4.1.1; The R Foundation for Statistical Computing, Vienna, Austria; http://www.R-project.org/). Continuous variables are presented as mean ± standard deviation, while categorical data are expressed as the number of participants (percentage of the total). Fisher’s exact test and t-test were used for univariate analyses to compare categorical and continuous variables, respectively. All statistical tests were two-sided; a P-value <0.05 was considered statistically significant. In the linear regression analysis, factors with a P-value <0.20 in the univariate analysis were included in the multivariate analysis to examine the clinical relationship of pre-specified clinical information with PI and ADI.

## Result

4D flow MRI was successfully performed, and measurements of PI at P-in, PI at P-out, and ADI were feasible for all 27 aneurysms. The values for PI at P-in and P-out were 0.95 ± 0.24 and 0.83 ± 0.22, respectively. Ultimately, the ADI in the study cohort was calculated to be 1.18 ± 0.28, which was significantly larger than 1.0 (P=0.0027 [t-test], **Figure 2**)

**Figure 2:**
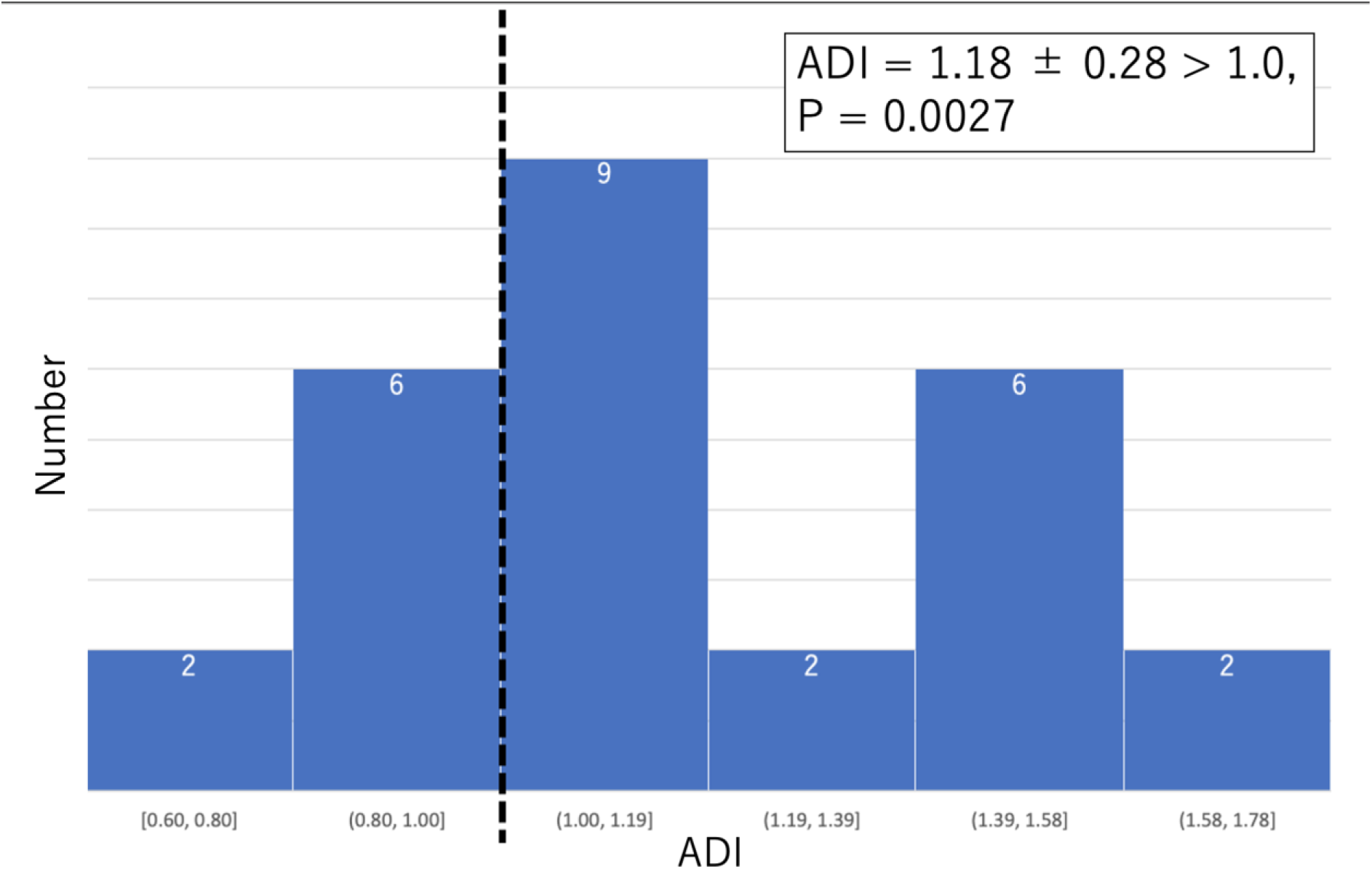
Histogram of the ADI on the aneurysm side. The mean ADI in the study group was 1.18, significantly exceeding 1.0 (P=0.0027, t-test).

A previous analysis showed that damping through the aneurysm could be observed with 4D flow MRI; however, the effect of the aneurysm on damping was not assessed. Upon comparing ADI with those of the contralateral side, it was found to be larger than the damping index of the contralateral normal artery (1.19 ± 0.30 vs 1.05 ± 0.17, P=0.029 [t-test], **Figure 3A and 3B**). However, there was a strong correlation between the damping index of the contralateral ICA and ADI (R^2^=0.94, **Figure 3C**). This analysis revealed that the damping effect of blood flow was greater in the cerebral aneurysms than in the corresponding normal arteries.

**Figure 3:**
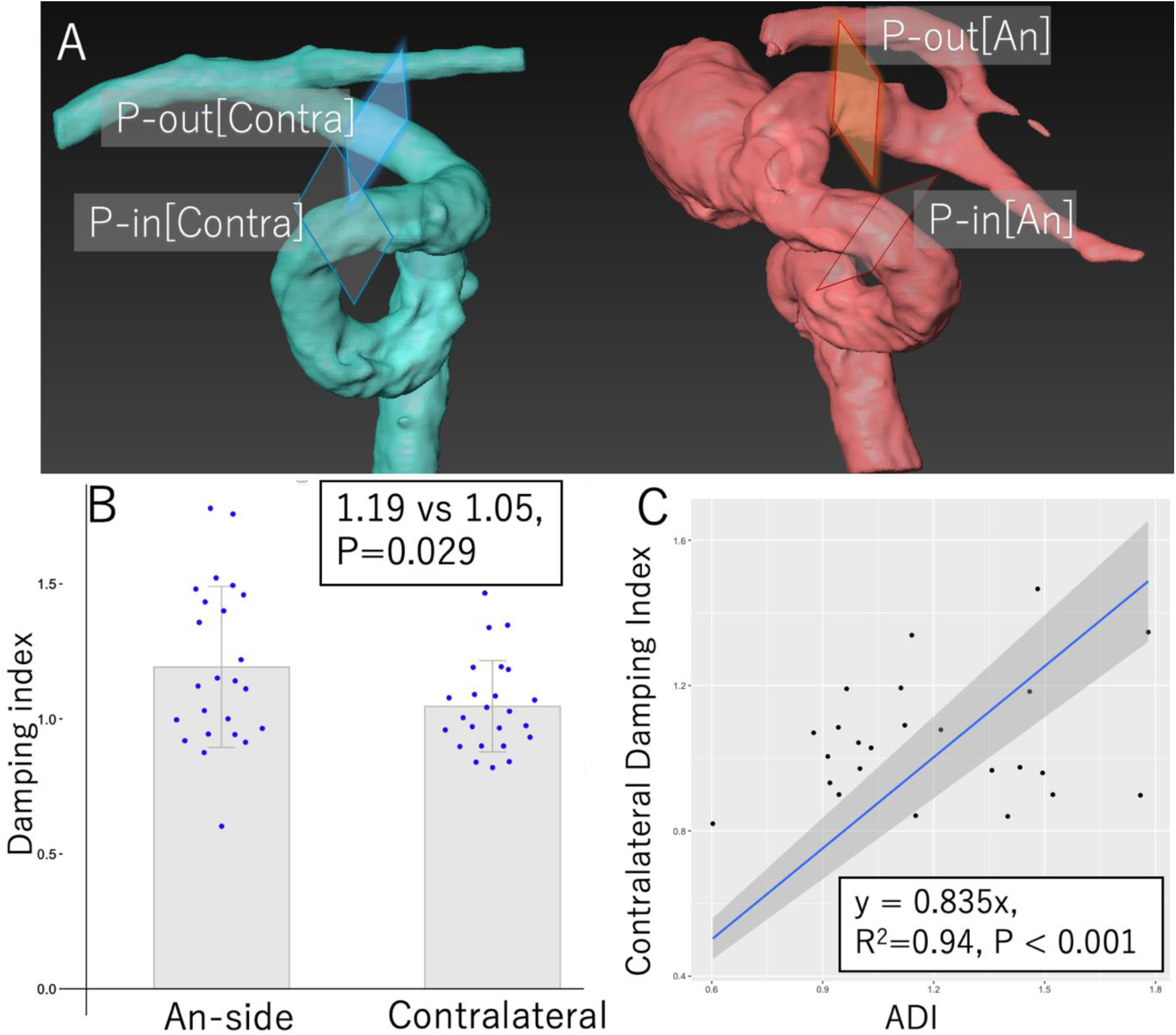
ADI comparison with the contralateral side. P-in and P-out on the contralateral side to the aneurysm are also defined at the symmetrical portion (Figure 3A). The ADI on the aneurysm side was significantly higher than that of the contralateral side (1.19 vs 1.05, P=0.029, t-test, Figure 3B). The ADIs on both sides exhibited strong correlations (R^2^=0.94, Figure 3C).

Regarding clinical factors related to PI at P-in, patients with a smoking history exhibited lower PI than patients without (1.07 ± 0.22 vs 0.81 ± 0.19, P=0.003 [t-test]). In the univariate regression analysis, PI at P-in showed a weak correlation with systolic blood pressure (P=0.013, R^2^=0.22), the grading of deep white matter hyperintensity on MRI fluid attenuation inversion recovery images (P=0.027, R^2^=0.18), and patient’s age (P=0.048, R^2^=0.16) (**Figure 4**). In a multiple regression analysis encompassing these factors, smoking history (β=-0.22, P=0.003), systolic blood pressure (β=0.005, P=0.032), and deep white matter hyperintensity grading (β=0.043, P=0.037) were identified as influencing factors of PI at P-in. This multivariate model explained 48.6% of the variation in the PI at P-in (R^2^=0.545, adjusted R^2^=0.486).

**Figure 4:**
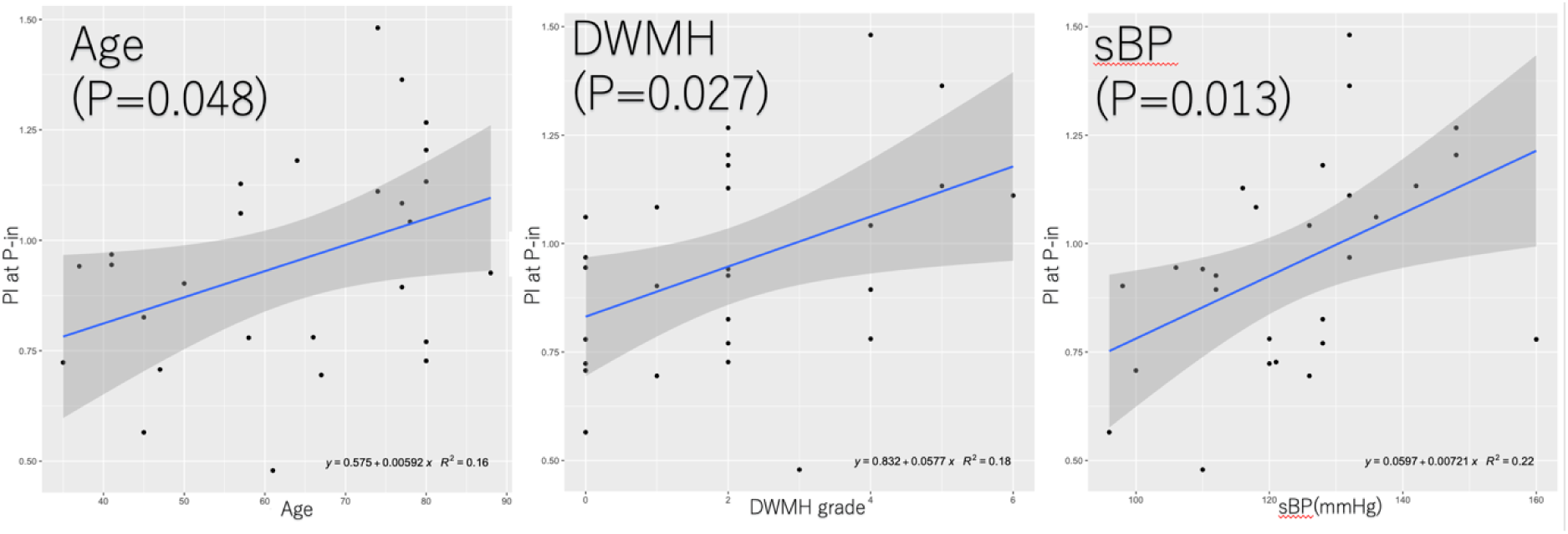
Factors related to PI at P-in on the aneurysm side. Age, deep white matter hyperintensity on MRI, and systolic blood pressure of the patients exhibit a positive correlation with their PI.

Regarding the clinical factors related to ADI, patients with a smoking history had a lower ADI compared to patients without (1.06 ± 0.26 vs 1.28 ± 0.27, P=0.045 [t-test], **Figure 5**). Although not statistically significant, patients on beta-blockers exhibited a higher ADI compared to patients without (1.60 ± 0.23 vs 1.15 ± 0.26, P=0.19 [t-test]). In a multivariate regression analysis including these two factors, both beta-blocker intake (β=0.46, P=0.015) and smoking history (β=-0.22, P=0.024) showed a statistically significant correlation with ADI. This model explained 28.4% of the variance in the ADI (R^2^=0.339, adjusted R^2^=0.284).

**Figure 5:**
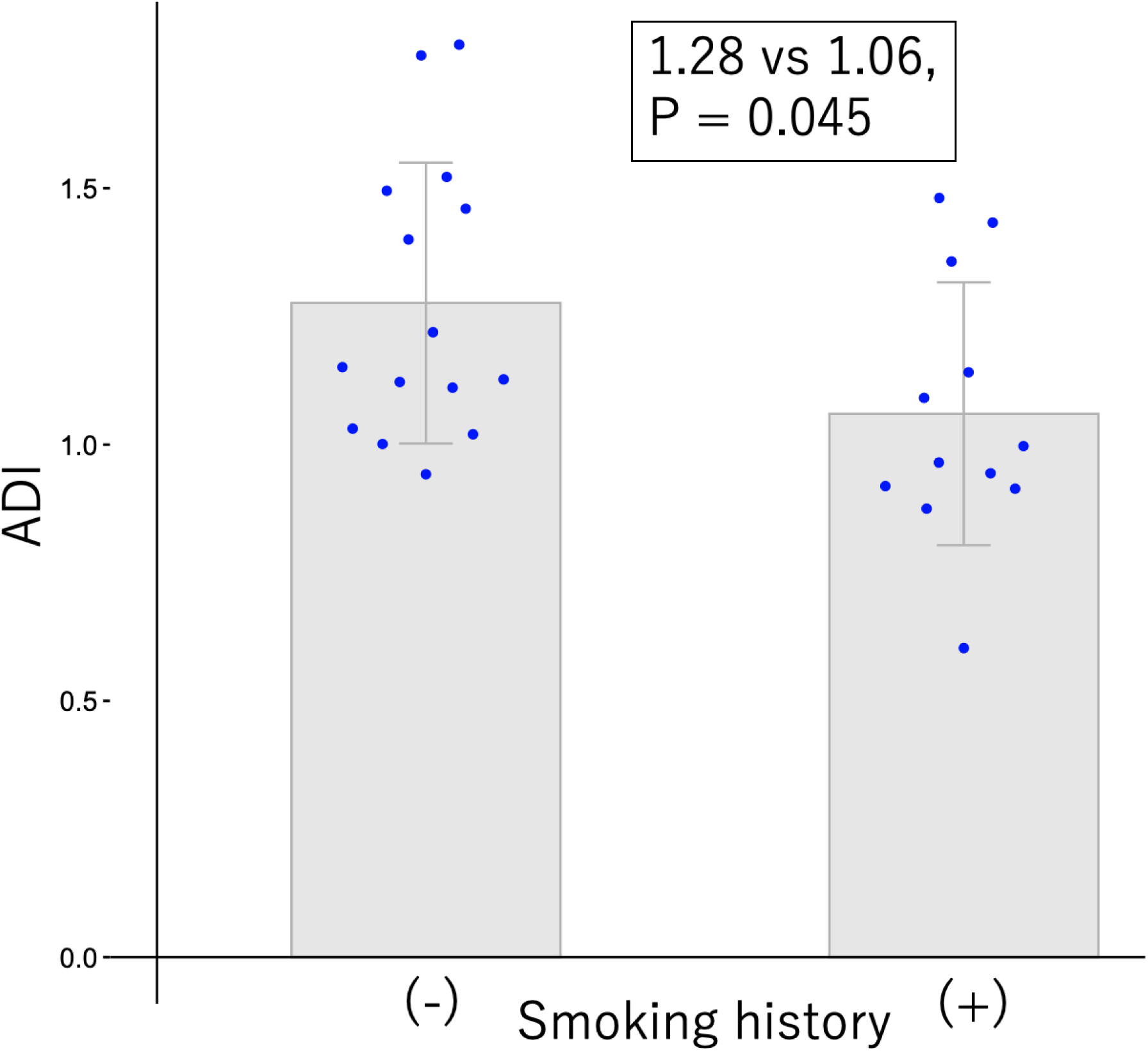
The relationship between smoking status and ADI. Patients with a smoking history have a significantly lower ADI than patients without.

## Discussion

In this study, we conducted an initial assessment of intracranial aneurysm stiffness using 4D flow MRI and successfully measured the pulsatile waveform within a cardiac cycle in the intracranial ICA and observed damping between the proximal and distal portions of the aneurysm in clinical cases with 3T MRI. We found that the measured extents of arterial pulsatility and blood flow waveform damping, indicated by PI and ADI, respectively, were associated with clinical factors such as smoking, blood pressure, antihypertensive drugs, and white matter hyperintensity on MRI.

We proposed ADI as an indicator of aneurysm rigidity. Previous studies have reported the measurement of PI in intracranial small vessels using 4D flow MRI with 7T machines and its association with clinical factors such as patient age and intracranial small vessel disease. ^8,9^ However, this is the first study to focus on measuring intracranial aneurysm stiffness. If the ADI is high, it means that the aneurysm has high compliance, and pulsatile energy is absorbed inside the aneurysm. Conversely, a low ADI suggests that the aneurysm wall is stiff, and the pulsatile waveform is conducted more directly to the arteries distal to the aneurysm. In normal large intracranial vessels, arterial damping acts as a buffer to protect the cerebral vasculature from pulsatile flow, but its role in pathological conditions such as intracranial aneurysms is scarcely reported. ^12,13,17^ In cardiovascular medicine, the stiffness of the central and peripheral arteries is well-documented as a biomarker for various cardiovascular events. ^2,3^ Similarly, the ADI assessment method proposed in this study may serve as a useful biomarker for predicting the natural history of unruptured intracranial aneurysms.

The association between arterial stiffness and clinical factors has been documented in the cardiovascular field. In extracranial arteries, antihypertensive drugs are associated with pulse wave velocity reduction, indicating a reduction in arterial stiffness. ^18,19^ Regarding beta-blockers, a possible explanation underlying this effect is the enhanced release of endothelium-derived nitric oxide, thereby improving endothelial function. ^20,21^ Similarly, the association between arterial stiffness and smoking is well-documented. Smoking increases arterial stiffness, which decreases after smoking cessation. ^22–24^ Chronic smoking is shown to be associated with reduced endothelial-dependent vasodilation, nitric oxide generation, and endothelial nitric oxide synthase activity.^25^ In the current study, consistent with previous finding regarding systemic arteries, ADI correlated positively with beta-blocker use and negatively with smoking. Theoretically, pulsatile blood flow collides directly with the rigid aneurysm wall in low-ADI aneurysms. In contrast, in high ADI situations, blood flow is cushioned by vessel wall compliance, and the impact on the aneurysm wall becomes milder. Consequently, a high ADI should protect against aneurysm growth and rupture. Future studies focusing on the association between clinical factors and ADI may lead to the development of optimal medical management strategies for stabilizing aneurysms.

From a clinical perspective, the physical properties of the intracranial arterial wall can affect the natural progression and treatment outcomes of intracranial aneurysms. Traditionally, morphological parameters such as aneurysm size and aspect ratio have been associated with the growth, rupture, and postoperative recurrence of unruptured intracranial aneurysms. ^26–29^ However, recent observational studies have revealed that the rate of aneurysm occlusion following flow-diverter stenting is influenced by patient factors such as hypertension, age, and smoking status. ^30–32^ In the era of novel treatment devices such as flow diverters, the properties of the vessel wall are crucial when considering their interaction with endovascular devices. Future studies on arterial stiffness will be vital for predicting long-term outcomes after intracranial aneurysm treatment or developing innovative treatment devices.

This study has a few limitations. First, it is a preliminary study with few patients from a single institution. Second, our proposed method estimates damping throughout the entire aneurysm volume and does not specifically focus on the vessel wall properties of a particular region within the aneurysm. As our group recently reported, blood flow analysis using 4D flow MRI can be useful for estimating the wall thickness of a specific region inside the aneurysm. ^33^ This approach should be considered a future direction for 4D flow MRI blood flow analysis. As this study only presents preliminary results, further studies are needed to explore the usefulness of assessing vessel wall properties using 4D flow MRI in basic research and clinical applications.

In conclusion, we proposed a novel method for assessing intracranial aneurysm wall stiffness using 4D flow MRI. Damping of the arterial pulse wave through the aneurysm can be observed, and PI and ADI were correlated with clinical factors, such as blood pressure, white matter hyperintensity on MRI, and smoking. Further studies are needed to explore less invasive assessment of aneurysm stiffness and its clinical applications.

## Data Availability

The datasets generated and/or analyzed during the current study are available from the corresponding author on reasonable request.

## Acknowledgment

None

## Non-standard Abbreviations and Acronyms

ADI: Aneurysm Damping Index
MRI: Magnetic Resonance Imaging
PI: Pulsatility Index
FOV: Field of View
TOF-MRA: Time-of-Flight Magnetic Resonance Angiography
TR: Repetition Time
TE: Echo Time

